# Physical Restraint Use in a US Intensive Care Setting – Protocol for a retrospective cross-sectional analysis using the MIMIC-IV repository

**DOI:** 10.1101/2025.04.23.25326294

**Authors:** Maximin Lange, Leo A Celi, Ben Carter, Jesse D Raffa, Sharon C O’Donoghue, Tom J Pollard

**Affiliations:** Laboratory for Computational Physiology, Harvard-MIT Division of Health Sciences and Technology, Institute for Medical Engineering and Science, Massachusetts Institute of Technology, Cambridge, MA, USA; Department of Psychosis Studies, Institute of Psychiatry, Psychology & Neuroscience, King’s College London, London, UK; Beth Israel Deaconess Medical Center, Boston, MA, USA; Department of Biostatistics, Harvard T H Chan School of Public Health, Boston, MA, USA; Department of Biostatistics and Health Informatics, Institute of Psychiatry, Psychology & Neuroscience, King’s College London, London, UK; Beth Israel Deaconess Medical Center (Retired), Boston, MA, USA

## Abstract

**Objective:** To investigate disparities in physical restraint use in a U.S. intensive care unit (ICU) setting, focusing on the influence of demographic factors (ethnicity, sex, age), mental health diagnoses, intubation status, and ICU type. The study also examines trends before and after policy changes in 2014.

**Methods:** This retrospective cross-sectional study uses MIMIC-IV data from adult ICU patients (2008–2022) at Beth Israel Deaconess Medical Center. The primary outcome is the proportion of ICU days with physical restraint. A binomial Generalized Linear Model (GLM) with a logit link function will be used to estimate associations between patient factors and the proportion of ICU time spent in restraints, modeling the number of days with restraint as a binomial outcome with the number of trials equal to the total ICU length of stay. Results will be reported as adjusted odds ratios with 95% confidence intervals. Temporal trends will be evaluated across predefined three-year intervals. Secondary analyses include binary restraint use (yes/no), death within 24 hours of restraint use (yes/no), interaction effects, and multiple sensitivity analyses.

## 1 Background

Physical restraints in healthcare settings serve as a safety measure for both patient and clinical staff [1, 2]. They may only be used when patients present an immediate danger to themselves or others, and after exhausting alternatives [3]. Risks from restraints are severe and potentially lethal, including asphyxiation, aspiration, blood clots, trauma, and mortality from any cause [4, 5, 6, 7, 8]. Restraints can further lead to psychological harm, causing mental health deterioration and reduced trust in healthcare, which may result in patients skipping follow-up care [9]. A number of professional and academic institutions have started to recommend a reduction of restraint use across all patient populations [1, 10, 11]. In 2014, Form CMS-10455 [12] was introduced, which required hospitals to report deaths associated with restraint and/or seclusion.

There have been a number of epidemiological studies regarding frequency of restraint utilisation. In recent years, an increasing number of studies have focused on understanding disparities in ethnicity, sex, gender and consequential risk of restraint [13, 14, 15, 16, 17]. Most studies on the topic have been conducted in an emergency department setting.

These studies come to conflicting conclusions – some find evidence for certain ethnicities to be restrained more than others, while some do not – yet, a meta-analysis [18] concluded that Black patients were at a higher risk of being physically restrained in emergency departments in relation to other ethnic groups, while Hispanic patients were less likely to be restrained compared with non-Hispanic patients.

Research on restraint use in ICUs highlights significant variations in prevalence and associated risks. A multicenter study [19] found that the prevalence of physical restraint use in acute-care hospital settings was as low as 8.7%. In critical care units, a separate study reported a 19.11% overall prevalence, increasing to 42.10% in patients with endotracheal tubes [20]. Others [21] report that 53% of mechanically ventilated patients on Canadian ICUs were physically restrained, with an average restraint duration of just over 4 days. This is very high compared to emergency department physical restraint utilisation rates of around 1% [18].

Factors associated with restraint use include sedation practices, agitation levels, and institutional policies, with some studies suggesting that restraints are applied inconsistently across patient populations rather than purely based on clinical necessity [22, 23]. Efforts to minimize restraints have included nonpharmacologic interventions, staff training, and standardized protocols, but adherence remains inconsistent due to risk-averse cultures and variations in institutional commitment [24].

Reviews on restraint use in the ICU [25, 26] criticize the widespread use of physical restraints in ICUs due to their harmful consequences and the lack of strong evidence supporting their effectiveness.

Unlike emergency departments, which focus on acute assessment and stabilization with typically brief patient encounters, ICUs provide continuous critical care for extended periods to medically unstable patients. This fundamental difference in clinical context—including longer patient stays, higher acuity of illness, greater presence of life-sustaining equipment, and differing staffing ratios—may impact restraint decision-making and utilization patterns. Additionally, ICU patients frequently experience altered consciousness due to sedation, mechanical ventilation, or critical illness itself, presenting unique considerations for restraint use that differ substantially from the predominantly alert population typically seen in EDs.

Since Minnick [27], who looked at ICU restraint patterns in a 2003-2005 time frame, limited numbers of studies have been conducted regarding prevalence of ICU restraint use in the US. To our knowledge, there also appears to be insufficient investigations into the influence of demographic attributes on frequency of restraint in a US ICU setting.

Understanding the patterns and disparities in ICU restraint use is crucial, as restraints pose significant physical and psychological risks, yet their application remains widespread and poorly regulated. While prior research has highlighted racial and demographic disparities in restraint use within emergency departments, few studies have examined whether similar biases exist in ICUs, where patients experience prolonged critical care. If restraint use is found to be influenced by demographic factors rather than purely clinical necessity, this raises ethical concerns about implicit bias in critical care decision-making. Identifying such disparities could inform institutional policies, improve staff training, and promote alternative strategies that reduce reliance on restraints, ultimately enhancing patient safety and equity in ICU settings.

## 2 Study Aim and Objectives

This study investigates disparities in physical restraint use across different demographic groups in the intensive care setting.

### Specific objectives

1. To determine whether ethnicity is independently associated with physical restraint use and duration in ICU patients
2. To assess the impact of other demographic factors (age, sex) on restraint use and duration
3. To examine whether the presence of specific mental health diagnoses affects the likelihood and duration of restraint application
4. To examine whether intubation status or the type of ICU affects the likelihood and duration of restraint application
5. To evaluate temporal trends in restraint use and duration before and after policy changes in 2014

## 3 Methods

### 3.1 Study Design

Retrospective cross-sectional study using electronic health records.

### 3.2 Setting

MIMIC-IV [28] contains data from patients who were admitted to the Beth Israel Deaconess Medical Center (BIDMC) in Boston, MA, United States.

### 3.3 Population

The study population will include all unique ICU patients aged ≥ 18 available on the MIMIC-IV electronic health records database from a 14-year period between 2008 and 2022, with any chief complaint. Multiple visits during the period by the same individual will be capped and only the first visit be considered.

### 3.4 Inclusion Criteria

- Adult patients (age ≥ 18 years) admitted to any ICU unit (Medical ICU, Surgical ICU, Cardiac ICU, etc.) for a minimum duration of 24h
- Admission between 2008 and 2022

### 3.5 Restraint Group

Restraint days will be identified from clinical notes and nursing documentation containing mentions of physical restraint, four-point restraint (including variations ‘4-point’), wrist restraint, soft restraint, leather restraint, or the ICD9 or ICD10 code for physical restraint utilization (V4987 & Z781, respectively).

Otherwise, patients will belong to the ‘non-restraint’ group.

### 3.6 Outcomes

#### 3.6.1 Primary Outcome

Proportion of ICU days with physical restraint use at patient level calculated as:

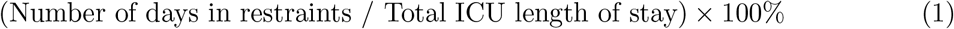

A “restraint-documented day” will be defined as any unique calendar date during the ICU stay on which at least one chart event matching the restraint keywords occurred.

#### 3.6.2 Secondary Outcome

- Restrain utilisation, yes or no
- Death occurred within 24h of restraint, yes or no

### 3.7 Statistical Analysis

#### 3.7.1 Descriptive Statistics

Descriptive statistics will be presented for all variables. Continuous variables will be reported as means with standard deviations or medians with interquartile ranges depending on their distribution. Categorical variables will be presented as frequencies and percentages.

#### 3.7.2 Primary Statistical Model

To analyze the primary outcome (proportion of ICU days with physical restraint), we will employ a binomial Generalized Linear Model (GLM) with a logit link function. For each patient, we will model the number of days with restraint as a binomial outcome with the number of trials equal to their total ICU length of stay.

The model will include the following predictors, all collected at day 1:

- Ethnicity (primary variable of interest)
- Age (as a continuous variable)
- Sex (male/female)
- Intubation status (yes/no)
- Presence of lines on body (central/arterial/dialysis/EVD/ECMO)
- Severity of Illness (SOFA, OASIS, SAPSII)
- Non-invasive ventilation mask (yes/no)
- English proficiency (yes/no)
- Level of agitation and sedation (RASS/CAM)
- Attempted suicide (yes/no)
- Chemical restraint received (haloperidol, droperidol, ziprasidone, olanzapine, ketamine, propofol, benzodiazepine, oxazepam, temazepam, clonazepam, alprazolam, midazolam, lorazepam, diazepam, dexmedetomidin, diphenhydramine)
- Time period (categorized in three-year intervals: 2008-2010, 2011-2013, 2014-2016, 2017-2019, 2020-2022)
- ICU type (Medical, Surgical, Cardiac, etc.)
- Path of admission (emergency, elective, transfer)
- Pain scale, regular, CPOT Scale
- Alcohol (CIWA/CIWA-AR)
- Presence of mental health diagnoses

Mental disorder categories include:

- Psychosis spectrum disorder (ICD10 F2’, ICD9 295’), (yes/no)
- Mania and bipolar disorder (ICD10 F30’, F31’, ICD9 296’), (yes/no)
- Substance use (ICD10 F1’, ICD9 303’, 304’), (yes/no)
- Mental disorder due to physiological conditions (ICD10 F0’, ICD9 293’), (yes/no)

Results will be presented as adjusted odds ratios with 95% confidence intervals and p-values, with significance set at 0.05.

#### 3.7.3 Model Selection

Model selection will use forward stepwise selection. Starting from a minimal model, we will add clinically meaningful variables one at a time based on expert consultation and prior literature. Variables will be retained if they meet statistical significance (p*<*0.05) and improve model fit as measured by AIC and the Hosmer-Lemeshow goodness-of-fit test. Two-way interaction terms between ethnicity and other covariates will be tested in a secondary step using the same criteria.

#### 3.7.4 Secondary Analysis

For the secondary outcomes—binary restraint utilization (yes/no) and death occurring within 24 hours of restraint (yes/no)—we will employ logistic regression using the same set of predictor variables. This will allow us to examine factors associated with the decision to use restraints at all, as well as potential short-term mortality following restraint use.

#### 3.7.5 Population and Multiple Visits

We will include only the first ICU visit for each unique patient within the study period to maintain independence of observations.

#### 3.7.6 Interaction Effects

As an exploratory analysis, we will assess two-way interaction terms between ethnicity and key variables (mental health diagnoses, intubation status, and time period) to explore whether disparities in restraint use differ by clinical context. Interaction terms will be added individually and retained based on likelihood ratio tests and AIC comparisons to evaluate model improvement.

#### 3.7.7 Sensitivity Analyses

Several sensitivity analyses will be conducted. First, we will stratify the analysis by ICU type to account for variation in restraint practices across different unit types. Second, we will perform a complete case analysis, including only patients with no missing data, to assess the potential impact of missingness. Third, we will conduct a propensity score matched analysis treating ethnicity as the exposure. Propensity scores will be estimated using logistic regression based on age, sex, ICU type, and other covariates. Matching will be restricted to Caucasian and African American patients, as these are expected to be the largest subgroups. The binomial GLM will then be re-estimated in the matched cohort, using the same variable selection approach as in the primary analysis. Lastly, we will repeat the analysis using alternative definitions of restraint use based on varying documentation patterns to evaluate the robustness of our findings.

#### 3.7.8 Temporal Trend Analysis

To examine temporal trends in restraint use, we will calculate the proportion of ICU days with restraint use for each three-year interval across the study period. The Cochran-Armitage test for trend will be used to assess the statistical significance of changes in proportions over time. Time interval will also be included as a categorical variable in the primary GLM to estimate adjusted changes in restraint use over time. We will test for interactions between time interval and ethnicity to evaluate whether demographic disparities have changed across time. Temporal trends will be visualized using time series plots stratified by key demographic characteristics.

#### 3.7.9 Model Diagnostics

We will assess model fit using standard diagnostics for GLMs including deviance and Pearson residuals, Cook’s distance for influential observations, and the Hosmer-Lemeshow goodness-of-fit test. Variance inflation factors will be calculated to check for multicollinearity among predictors. We will also examine calibration plots to assess the agreement between observed and predicted proportions.

If any of the variables are too sparsely populated, some groups might need to be collapsed.

#### 3.7.10 Sample Size Considerations

No a priori power calculation was performed as this is a retrospective analysis using all available data from the MIMIC-IV database. With data from over 40,000 ICU stays, the sample size is expected to be sufficient to detect clinically meaningful differences in restraint use across demographic groups.

#### 3.7.11 Software

All analyses will be conducted using Python in Google Colab.

### 3.8 Data availability

Source code for SQL extraction code for the MIMIC-IV database will be made public on GitHub, along with all python code used for data analysis. MIMIC data is available on PhysioNet after completion of relevant research training and accreditation.

## 4 Ethics

This research will be conducted in accordance with the Declaration of Helsinki and follow international ethical standards for medical research involving human data.

MIMIC-IV data was collected as part of routine clinical care. It has been deidentified and transformed. It is available to researchers who have completed training in human research and signed a data use agreement. It was approved for research by the institutional review boards of the Massachusetts Institute of Technology and BIDMC, who granted a waiver of informed consent and approved the sharing of the research resource. Individual patient consent will not be required for publication as this was addressed in the original approval for the MIMIC-IV databases, which permits research use and publication with proper de-identification maintained.

## 5 Contributors

LAC conceived the idea of investigating intensive care physical restraint prevalence. BC, JDR and ML developed the statistical analysis plan which was checked by TJP and LAC. ML and TJP wrote the first draft of the protocol. LAC and SOD provided clinical guidance. TJP, BC, JDR, SOD, and LAC offered supervision, read and commented on the protocol.

## Notes

### Competing Interest Statement

The authors have declared no competing interest.

### Funding Statement

No specific funding was obtained for the delivery of this study. Maximin Lange is supported through a studentship by the London Interdisciplinary Social Science Doctoral Training Partnership (LISS DTP). Ben Carter is part funded through the NIHR Maudsley Biomedical Research Centre at the South London and Maudsley NHS Foundation Trust in partnership with Kings College London This paper represents independent research funded by the NIHR Maudsley Biomedical Research Centre at South London and Maudsley NHS Foundation Trust and Kings College London. The views expressed are those of the authors and not necessarily those of the NIHR or the Department of Health and Social Care. Leo Anthony Cell is funded by DS-I Africa U54 TW012043-01 and Bridge2AI OT2OD032701 and the National Science Foundation through ITEST #2148451. Tom Pollard is funded by NIH Bridge2AI OT2OD032701 and RO1 EB030362. Jesse Raffa is funded by Philips Healthcare. The funders had no role in study design, data collection, analysis, decision to publish or preparation of the manuscript.

### Author Declarations

MIMIC-IV data was collected as part of routine clinical care. It has been deidentified and transformed. It is available to researchers who have completed training in human research and signed a data use agreement. It was approved for research by the institutional review boards of the Massachusetts Institute of Technology and BIDMC, who granted a waiver of informed consent and approved the sharing of the research resource.

